# Determinants of Tuberculosis Diagnosis Delays among Patients Attending Public Health Facility of Metekel Zone, Northwest Ethiopia, 2024 unmatched Case- Control Study

**DOI:** 10.64898/2026.11.17.26368197

**Authors:** Aboma Tolessa Denta, Andarge Bayro Burusso, Teklu Nega Yimenu

## Abstract

**Background:** Tuberculosis Diagnosis delays among tuberculosis patients can worsen infection transmission, disease severity, and mortality risk. Identifying factors influencing delays is crucial due to its persistence across healthcare levels and communities. The study identify determinants of tuberculosis diagnosis delay among patients attending public health facility of Metekel zone, Northwest Ethiopia:2023

**Methods:** An unmatched case-control study was conducted in Metekel zone health facilities from March to April 2023, involving 210 tuberculosis patients (103 cases, 107 controls) selected through systematic sampling. Data collected through structured questionnaires and STATA used for analysis, including bi-variable and multi-variable logistic regressions. Variables with p-value < 0.25 in bi-variable analysis were included in the final model after checking model fitness and multicollinearity.

**Results:** Overall, 210 tuberculosis patients were included in the study. The following independent variables were associated diagnosis delay of tuberculosis: TB patients who travel greater than 10km (AOR: 2.941, (95 % CI: 1.509, 5.731); TB patients who are living at rural area (AOR: 4.135, 95 % CI :1.759, 9.723); TB patients who prior visit in holly-water and traditional medicine [AOR: 2.978, (95% CI: 1.292, 6.863); poor knowledge about TB [AOR: 2.826, (95% CI: 1.404, 5.687)].

**Conclusion:** Tuberculosis diagnosis delay is a major challenge to eliminate tuberculosis by breaking the chain of transmission. This study can be attributed to factors including place of residence being rural, poor knowledge about tuberculosis, non-health facility initial actions upon symptom onset, and far distance to health institutions. Recognizing these determinants helps in identifying and addressing barriers inhibiting early diagnosis, potentially improving healthcare access and promoting more timely diagnosis and treatment for tuberculosis patients.

## INTRODUCTION

## BACKGROUND

Diagnosis delay of tuberculosis is the time interval between the beginning of symptoms and confirmation of tuberculosis in the patient (1,2). Diagnosis delay in tuberculosis, and early initiation of treatment results in augmented rate of infectivity in the society, and it is estimated that untreated smear positive patients can infect on average 10 contacts annually and 20 during the natural history of the disease until death(3). Delayed diagnoses occur when a patient seeks medical treatment for symptoms, and yet the condition causing those symptoms goes undiagnosed for an unreasonable amount of time. Often, this may result in the condition worsening into a serious illness or disease(4). Delays in diagnosis can directly impact a patient’s chance of recovery and survival. The earlier an illness is correctly diagnosed and treated, the better chance the patient has at recovery and survival. The failure to diagnose health issues early can lead to more serious complications of the illness or even death(5).

In 2021, only 6.4 million from the estimated 10.6 million people who developed active TB were diagnosed and notified(6). A WHO-recommended rapid molecular test was used as the initial diagnostic test for only 38% of the 6.4 million people newly diagnosed with TB in 2021 (6, 7). The burden of drug-resistant TB (DR-TB) is also estimated to 450 000 new cases of rifampicin- resistant TB (RR-TB) in 2021(6). An estimated 2.5 million people fell ill with tuberculosis in the African Region in 2021,and as many as half-a-million people died due to the disease in the same year(8). Despite making up only 15% of the world’s population, the region accounted for 23% of new TB cases and 31% of TB-related deaths in 2021(6). According to the World Health Organization (WHO), in 2020, Africa accounted for approximately 25% of the estimated global TB cases. The impact of TB on African populations is influenced by socioeconomic factors, healthcare infrastructure, and access to diagnostics and treatment services(9).

Ethiopia remains among the global high TB burden countries, with an annual TB incidence of 119 cases per 100,000 population in 2021. The WHO estimates that about 30 percent of TB cases go undetected by the healthcare system in Ethiopia, resulting in unnecessary deaths(10). With an estimated 143, 000 new cases and 21,100 TB- related deaths in 2021, Ethiopia ranks fifteenth and seventh in the world and Africa region, respectively, among the countries with the highest tuberculosis burden(11) Ethiopia in 2020 among the estimated incident TB cases, total of 108,196 persons with all forms of TB were notified to the national program. About 9.9% of the notified TB cases in 2020 were children less than 14 years of age (12, 13).

Prompt diagnosis is critical for effective TB control program performance and achieving the End TB targets(14). The End TB Strategy has set ambitious milestones for 2025 and targets to end TB by 2035. Three pillars place precedence on expanding patient-centered TB prevention and control efforts, forming policies and multi-sector collaborations across communities and public and private sectors, and ensuring continued attention on scientific research innovations for TB care(14). Ethiopia has adopted the post-2015 TB strategy called the End TB Strategy which aims to end the TB epidemic by 2035 by reducing TB incidence by 90% and mortality by 95% from the 2015 baselines, and percent of affected households facing catastrophic costs due to TB to zero(15).

Most humans (about 90%) who develop the disease are adults, with greater cases among men than women. About a quarter of the world’s population has been infected with M. tuberculosis(16).Tuberculosis is the leading cause of deaths every year among the infectious disease worldwide alongside HIV. It kills more than five thousand children, women and men each day(17). Early diagnosis and appropriate treatment are integral to reduce the spread of tuberculosis (TB)(18). The advantageous control of the disease relies upon on the search for primary health care (PHC), through the patient, within the period of 2–3 weeks (19),(20) and the action of the PHC as a gateway for the services; facilitating the access and promoting full high- quality care; aiming at promoting, protecting, and recovering health (21).

Duration of tuberculosis diagnosis is the time interval between the onset of symptoms and confirmation of TB in the patient. The main tool for controlling tuberculosis (TB) is the early diagnosis and initiation of treatment for people affected by the disease. For this, early detection of respiratory symptoms (RS) is necessary, those people who have persistent cough for two or three weeks. The active search in the community then becomes an essential tool, strengthening the decentralization of TB control actions to primary health care (PHC)(22).

Diagnosis delay is the time interval from the onset of symptoms of TB until the first visit to any formal health care facility within 30 days as cut-off points. WHO classifies person who presents with symptoms or signs suggestive of TB most common a productive cough or unexplained cough with or without other respiratory or constitutional symptoms lasting 2-3 weeks as TB suspect and the patient will seek treatments within the given period, patients presented to modern health facility more than 2-3 weeks considered as patient delay as WHO recommendation. However, studies added extra margin of 7 days to take 30 days as cut-off to assess magnitude of patient delay: a delay >30 day(23). Diagnosis delay of tuberculosis (TB) can lead to an increased infectivity period, delayed treatment, and multiplied severity of the disease in our community.

## Methods

### Study design

A facility-based unmatched case-control study was conducted from March-5 to April 30, 2023.

### Study setting and period

Metekele Zone is located 546 km from Addis Ababa and 352km from Assosa, the capital city of the Benishagul Gumuz region of Ethiopia. For administrative purposes, the zone is structured into seven districts and a town Administration. In the study area, from 25 health facilities only 17 health facilities give TB diagnosis and DOT program. In the Metekele zone, all tuberculosis patients receiving anti-TB drugs at the healthcare facility providing tuberculosis diagnostic services were included in the study population. This conducted from march-05 to April 30,2023.

### Source and study Population

#### Source Population

All adult tuberculosis adult patients who took Anti-tuberculosis treatment at Metekele zone public health facilities.

#### Study population

The study population of cases were all adult tuberculosis patients, who present at selected a health facility greater than 21 days after the onset of the disease and confirmed tuberculosis diagnoses. while controls are adult tuberculosis patients, who present at a health facility before 21 days after the onset of the disease and confirmed tuberculosis diagnoses.

#### Selection of cases and control

Identification and selection of cases and controls was conducted at each health facility’s TB clinics from TB records and medical cards. To select the study subjects, a list of tuberculosis patients was prepared from the TB logbook. The TB patient’s medical cards were traced by card number and collected from the respective health facility. Those patients whose duration of the first presented symptom and confirmed tuberculosis diagnoses greater than 21 days were entered into the case group, whereas those patients whose duration of the first presented symptom and confirmed tuberculosis diagnoses was less than or equal to 21 days were entered into the control group. For interview purposes, both groups are listed separately.

### Inclusion criteria and Exclusion criteria

#### Inclusion criteria

All adult tuberculosis patients who took Anti-tuberculosis treatment at selected Metekele zone public health facilities in the registries from November-05 to April 30, 2023.

#### Exclusion criteria

The participants in the TB registry unit who were transferred from the study area during study the period: Those who were unable to communicate and seriously ill.

### Study Variables

#### Dependent variable

Diagnosis delay

#### Independent variables

Socio demographic: age, sex, education, marital status, religion, residence, Knowledge, Distance from health institution, Multiple health facility visits, Health seeking behavior, First visiting traditional health facility, Clinical characteristics of tuberculosis

### Operational definitions

**Diagnosis delay**: is defined as the time interval between the onset of symptoms tuberculosis and diagnosed as tuberculosis patient(2, 67).

**Cases:** the study subjects, or tuberculosis patients, who present at a health facility greater than 21 days after the onset of the disease and confirmed tuberculosis diagnoses (51) (67)

**Control**: those tuberculosis patients who visit a health facility in less than or equal to 21 days after the onset of the disease and confirmed tuberculosis diagnoses (68)

**Patient diagnosis delay**: The time interval between the onset of major symptoms of TB to the first diagnosis as tuberculosis patient within 21 days as cut off points. Hence Patients who presented at health care facilities more than 21days after the onset of the major symptoms of TB and diagnosed patients are considered as delayed based on WHO standard definition(69, 70).

**Assessment of knowledge about tuberculosis:** Patients’ knowledge was assessed by asking them 12 questions – questions on disease etiology, mode of transmission, and treatment and prevention of tuberculosis. A scoring system was generated to assess the level of knowledge. Each correct answer was awarded one point and each wrong answer was awarded zero. Those

who scored 50% and above were considered to have “good knowledge” while those who scored less than 50% were considered to have “poor knowledge” about tuberculosis(71).

**Healthcare seeking behavior:** is first action undertaken to the illness by individuals who observe a symptom of tuberculosis(72).

**The onset of tuberculosis symptom**: The time at which the first symptom (i.e. Cough and other constitutional symptoms like fever, weakness, and weight loss or chest pain) of the illness for which a patient’s health care seeking begun(73).

### Ethical consideration

Ethical clearance was obtained from the Ethical review committee of Debre Markose University, College of Health Science.

### Data Analysis

The collected data was entered into Epi-data version 4.6 and exported STATA for analysis. Associations between the dependent variable (diagnosis delay) and the independent variables were analyze by calculating the odds ratio. Data were summarized using frequency, mean with SD. Data analysis with, logistic regressions were done. Those variables with p value of less than

0.25 in bi-variable binary logistic regressions were entered in to the multivariable binary logistic regression model. Statistically significant association was declared at p value of less than 0.05. Model fit was check by Hosmer-Lemeshow (>0.05) goodness of fit and inter correlation of the independent variable were checked by multicollinearity (tolerance >0.1, VIF<10).

## RESULT

### Socio-demographic characteristics of the patients

Two hundred (with a response rate of 95%) gave complete responses. Greater than half (54.4%) of TB patients were married. On the other hand, more than two third (80%) of TB patients were Orthodox Christian follower. Moreover, 89(44.5%) TB patients were hadn’t formal education.

156 (78.5%) TB patients were rural dwellers. The mean (±SD) age of TB patients was 34.74(±10.944) years old (table .1).

**Table 1:** Socio-demographic characteristics of study participants.

| Characteristics | Control |  | Cases |  |
|---|---|---|---|---|
|  | **Frequency** | **%** | **Frequency** | **%** |
| Age category |  |  |  |  |
| **18-30 years** | 46 | **23** | **45** | **22.5** |
| **31-45 years** | **46** | **46** | **28** | **14** |
| **46-60 years** | **15** | **7.5** | 15 | **7.5** |
| >=60 years | **3** | **1.5** | **2** | **1** |
| Sex |  |  |  |  |
| **Male** | 77 | **38.5** | 55 | **27.5** |
| **Female** | 33 | **16.5** | 35 | **17.5** |
| Educational status of TB patients |  |  |  |  |
| **no formal education** | 47 | **34** | 42 | **36.5** |
| **primary education** | 32 | **16** | 29 | **14.5** |
| **Secondary and above** | 31 | **15.5** | 19 | **9.5** |
| Residence |  |  |  |  |
| **Urban** | 34 | **17** | 9 | **4.5** |
| **Rural** | 76 | **38** | 81 | **40.5** |
| Marital status |  |  |  |  |
| **Married** | 57 | **28.5** | 52 | **26** |
| **Single** | 40 | **20** | 29 | **14.5** |
| **Divorced** | 11 | **5.5** | 8 | **4** |
| **Widowed** | 2 | **1** | 1 | **0.5** |
| **Religion** |  |  |  |  |
| **Orthodox** | 92 | **46** | 68 | **34** |
| **Muslim** | 4 | **2** | 12 | **6** |
| **Protestant** | 14 | **7** | 7 | **3.5** |
| **Others** | 0 | **0** | 3 | **1.5** |

### Health seeking care and clinical Characteristics

According to the finding of this study 90(45%) of the tuberculosis patients were diagnosed by microscopy. On the other hand, 160 (80%) of tuberculosis patients first sought for treatment at health facility. Beside of this, only 18(20%) of tuberculosis patients were had a history of contact of TB patients before (table 2).

**Table 2:** Health seeking and clinical Characteristics of tuberculosis patients.

| **Characteristics** | **Control** |  | **Cases** |  |
|---|---|---|---|---|
|  | **Frequency** | **%** | **Frequency** | **%** |
| **First sought treatment** |  |  |  |  |
| **Health Facility** | 98 | **49** | 62 | **31** |
| **holy spring &traditional medicine** | 12 | **6** | 28 | **14** |
| **Co-existence of chronic diseases** |  |  |  |  |
| **No** | 100 | **50** | 83 | **41.5** |
| **Yes** | 10 | **5** | 7 | **3.5** |
| **Have history of previous TB patient contact** |  |  |  |  |
| **No** | 104 | **52** | 78 | **39** |
| **Yes** | 6 | **3** | 12 | **6** |
| **Distance** |  |  |  |  |
| **<10 km** | 42 | **21** | 69 | **34.5** |
| **>=10km** | 48 | **24** | 41 | **20.5** |
| **Functional status patients** |  |  |  |  |
| **Working** | 101 | **50.5** | 78 | **39** |
| **Ambulatory** | 5 | **2.5** | 12 | **6** |
| **Bed ridden** | 4 | **2** | 0 | **0** |
| **Methods of diagnosis** |  |  |  |  |
| **Microscope** | 53 | **26.5** | 37 | **18.5** |
| **Chest X-ray** | 52 | **26** | 39 | **19.5** |
| **Gen xpert** | 3 | **1.5** | 9 | **4.5** |
| **Others** | 2 | **1** | 5 | **2.5** |

### Knowledge and behavioral related characters

Nearly two-third (66%) of tuberculosis patients were had a good knowledge about the causative agent, transmission mechanism and prevention methods of tuberculosis. On the other hand, 144(72%) and 33(16.5%) of tuberculosis patients had a habit of alcohol drink and smoking behavior (table 3).

**Table 3:** Knowledge and behavioral related characteristics participants.

| **Characteristics** | **Control** |  | **Cases** |  |
|---|---|---|---|---|
|  | **Frequency** | **%** | **Frequency** | **%** |
| **Smoking status** |  |  |  |  |
| **No** | 92 | **46** | 75 | **37.5** |
| **Yes** | 18 | **9** | 15 | **7.5** |
| **Alcohol drinking status** |  |  |  |  |
| **No** | 32 | **16** | 24 | **12** |
| **Yes** | 78 | **39** | 66 | **33** |
| **Knowledge** |  |  |  |  |
| **poor knowledge** | 26 | **13** | 42 | **21** |
| **Good knowledge** | 84 | **42** | 48 | **24** |

From the 200 TB patients’ 66% (95% CL: 59.5%, 72.5%) of patients had a good knowledge about tuberculosis disease (about causative agent, transmission and treatment as well as prevention mechanism). From the knowledge assessment questions the highest score 162(81%) of patients were aware about appetite loose is a typical sign and symptom of TB. Similar 161(80.5%) of patients also, know that night sweeting is the symptom of tuberculosis disease. Lastly, only 37(18.5%) of patients also respond that the treatment of tuberculosis disease is in charge (Figure 1).

**Figure 1.**
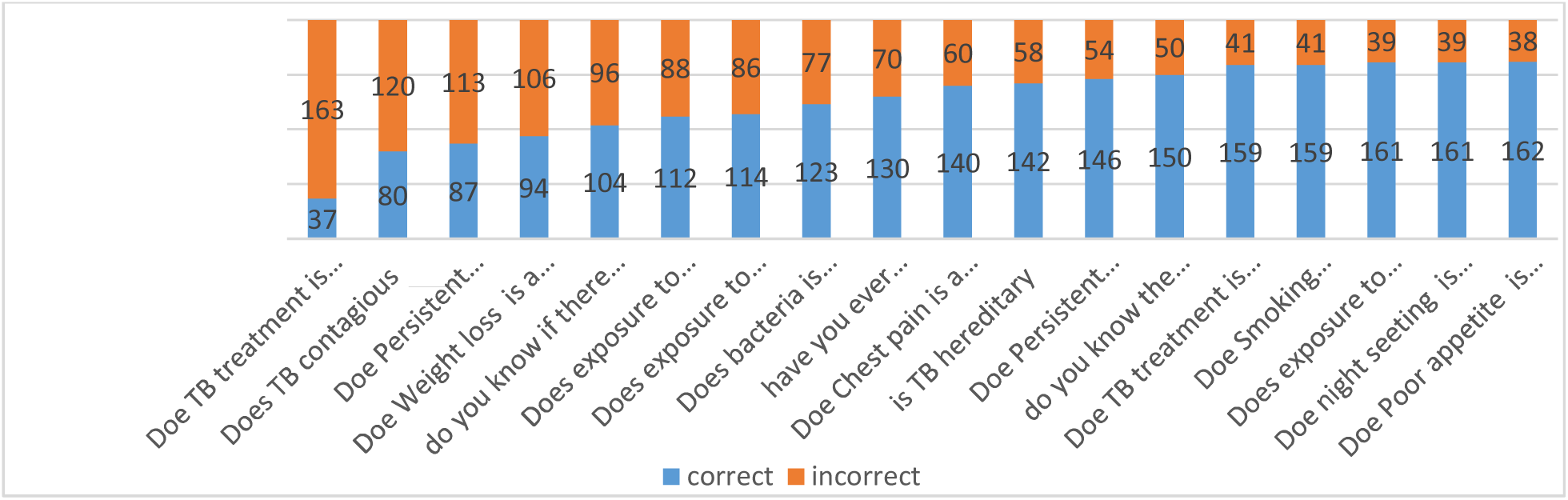
Patients responding about knowledge of tuberculosis at Metekel zone, north-west Ethiopia, 2023 (n=200).

### Factors significantly associated with tuberculosis diagnosis delays

During the bi-variable logistic regression analysis; knowledge of tuberculosis, Age, sought treatment before the first visit at public health facilities, educational status, residence, sex, alcohol drink, cigarette smoking, and type of diagnosis were found to be candidate variable for multivariable analysis (p-value ≤0.25). During the multivariable logistic regression analysis knowledge of tuberculosis, sought care before the first visit at public health facilities, residence and distance were significantly associated with delay in diagnosis of tuberculosis of tuberculosis.

The odds of diagnosis delay of tuberculosis among TB patients for those who travel greater than 10km were 2.9 times [AOR: 2.941, (95% CI: 1.509, 5.731)] higher than TB patients who travel less than 10km. Similarly, tuberculosis patients who are living at rural area were 4 times [AOR: 4.135, (95% CI: 1.759, 9.723)] more likely to delay than the urban dwellers. Additionally, TB patients who were attending treatment in holly-water and traditional medicine were 2.8 times more likely delay [AOR: 2.978, (95% CI: 1.292, 6.863)] higher than counterparts. The odds of diagnosis delay of tuberculosis among TB patients for those who had poor knowledge of tuberculosis were 2.8 times [AOR: 2.826, (95% CI: 1.404, 5.687)] higher than those patients who had good knowledge of tuberculosis (Table 7).

**Table 7:** Multivariable binary regression analysis of factors associated with delayed in diagnosis of tuberculosis among tuberculosis patients at Metekel zone, north-west Ethiopia, 2023.

|  | **Case** | **Control** | **COR** | **AOR** | **p-value** |
|---|---|---|---|---|---|
| **Age category of patients** |  |  |  |  |  |
| **18-30 years** | 46 | 45 | **1** | **1** |  |
| **31-45 years** | 46 | 28 | 0.622(0.333,1.16) | 1.084(0.119,9.895) | 0.943 |
| **46-60 years** | 15 | 15 | 1.022(0.448,2.333) | 0.732(.0080,6.676) | 0.782 |
| **>=60years** | 3 | 2 | 0.681(0.109,4.273) | 1.009(0.0101,10085) | 0.994 |
| **Chronic disease** |  |  |  |  |  |
| **Yes** | 7 | 10 | 0.843(0.308,2.313) | 0.690(0.212, 2.243) | 0.537 |
| **No** | 83 | 100 | 1 | 1 |  |
| **Health care seeking** |  |  |  |  |  |
| **Holy-spring &traditional medicine** | 28 | 12 | 3.688(1.747,7.787) | 2.978(1.292, 6.863) * | 0.01 |
| **Health Facility** | 62 | 98 | 1 | 1 |  |
| **Smoking status** |  |  |  |  |  |
| **Yes** | 15 | 18 | 1.022(0.483,2.164) | 0.478(0.193,1.180) | 0.056 |
| **No** | 75 | 92 | 1 | 1 |  |
| **Educational status** |  |  |  |  |  |
| **no formal education** | 47 | 42 | 0.296(.719,2.956) | 1.292(0.520,3.211) | 0.659 |
| **Primary education** | 32 | 29 | 1.479(0.691,3.164) | 1.502(0.617,3.657) | 0.375 |
| **Secondary and above** | 31 | 19 | **1** | **1** |  |
| **Sex** |  |  |  |  |  |
| **Male** | 55 | 77 | 0.673(0.374,1.213) | 0.571(0.294, 1.107) | 0.097 |
| **Female** | 35 | 33 | **1** | **1** |  |
| **Type of diagnosis** |  |  |  |  |  |
| **Microscopy** | 53 | 37 | 1.332(0.759,2.337) | 0.598(0.308, 1.159) | 0.128 |
| **Other method** | 57 | 53 | **1** | **1** |  |
| **Knowledge** |  |  |  |  |  |
| **Good knowledge** | 84 | 48 | 1 | 1 |  |
| **Poor knowledge** | 26 | 42 | 2.827(1.545,5.172) | 2.826(1.404,5.687) * | 0.004 |
| **Alcohol drink** |  |  |  |  |  |
| **Yes** | 78 | 66 | 1.28(0.605, 2.103) | 1.300(0.624, 2.705) | 0.483 |
| **No** | 32 | 24 | 1 | 1 |  |
| **Residence** |  |  |  |  |  |
| **Urban** | 34 | 9 | 1 | 1 |  |
| **Rural** | 76 | 81 | 4.026(1.812,8.948) | 4.135(1.759, 9.723) * | 0.001 |
| **Distance** |  |  |  |  |  |
| **<10km** | 69 | 42 | 1 | 1 |  |
| **>=10 km** | 41 | 48 | 1.923(1.092,3.389) | 2.941(1.759,5.731) * | 0.002 |

AOR: Adjusted odds ratio, COR: Crude odds ratio      \* Significant at p<0.05

## DISCUSSION

In this study tuberculosis diagnosis delays have been associated with various socioeconomic factories. The present study also revealed residence, as independent predictors of patient delay in TB diagnosis that means the patients’ who live dwell the rural residence more likely delay than tuberculosis patients who live the urban residence. This could be explained by the fact that study participants may have less adequate access to healthcare services and information if they live in a rural area as opposed to an urban one. Rural residents may not have the same oversight of healthcare workers as those living in urban areas, which could lead to a lower level of TB education(74). The possible suggestion for differences in delay rural residences could be probably by culture, low socioeconomic status, and lack of information about the availability of free tuberculosis diagnosis, treatment services and absence of frontline diagnostic services in rural settings. This finding supported different studies in southern Ethiopia Hadiya zone public health facilities residence is the significant factors for patient delays(66). The study support in Ethiopia patient delay is significantly different with residence (p<0.001), TB patients from rural longer patient delay for diagnosis of tuberculosis than those from urban(45). This study in line with study carried out Cambodia(75) showed that significant factors for diagnosis of tuberculosis between urban and rural areas were as rural Cambodia in delayed TB diagnosis. The other studies consistent with this study in Ahvaz rural residence were the most strong factors associated with diagnostic delays(76).

Regarding use of traditional tuberculosis care as the first choice had a strong effect on tuberculosis patients to access formal diagnostic source at the right time. As demonstrated in this study regarding health care seeking or first visiting tuberculosis patient who firstly visiting holly sprite or traditional medicine were more than two folds more likely to delay tuberculosis patient who visit health facility for tuberculosis diagnosis. This might cause low reliance of diagnosis in health facility and strong cultural bond in holy water and traditional healer treatments leading the respondents to seek care for diagnosis tuberculosis after failure of treatments in informal sources. In addition to in Metekele zone where there are various traditional practices exposures and disparities diagnostic access health facility, patient sough care from informal provider; as a result, the patients might have been given inappropriate care which led to several other visits before reaching the appropriate health facility for tuberculosis care. The other reason due to low awareness about the disease, about the severity and about availability of services, patients ignore TB symptoms and remain at home or visit holly water and traditional healers. In line with a previous study in Kampala, Uganda(77), this study shows that a significant proportion of TB patients used alternative healthcare as the first choice of care in Hohoe Municipality, with nearly 40 % initially presenting to traditional healers (78). Qualitative study which is highly consistent with this study carried out in Tanzania, one-third of patients first visited a traditional healer after the onset of symptoms, making this the place preferred as the first point of call by most patients. Patients tended to stay with the traditional healer for a long time. The delays in patients first visiting a traditional healer was almost three times as long as the delay in those first consulting a conventional health facility(56). This may be beliefs concerning the cause of the ailment was identified as an important determinant of first action health-seeking behavior.

In this study, accessibility to health care institution was measured by physical distance in kilometers from the patients’ residence to the nearest public health facility that provided the tuberculosis diagnostic service. Distance has been shown to significantly associate with a delay in diagnosis of tuberculosis. Patients who traveled greater than ten kilometers were more likely to delay than patients who lived within 10 kilometers radius. This finding suggests that increased travel distance may lead to prolonged periods before seeking medical care, possibly due to additional travel time, transportation challenges, and reduced access to immediate medical intervention. Patients living farther from healthcare facilities may face obstacles in promptly accessing diagnostic services, ultimately leading to delays in diagnosis. In the context of this study the associated due to the fact that scattered settlement of some population surrounding the catchment area, constraints of outreach and mobile health service and inaccessibility of diagnostic facility. This finding supports study in Ethiopia Jimma (79) have revealed that far walking distance between health facilities patients dwell was a significant factors for delays diagnosis of tuberculosis. Participants who lived beyond 10 Km radius from health facilities delay for diagnosis of tuberculosis. The other finding support this study was carried out in Ethiopia north shoa diagnosis delay (44),Amhara region Oromia special zone(60).

It is necessary to have adequate knowledge of tuberculosis in order to diagnosis tuberculosis help as soon as possible. In this study tuberculosis patients who had poor knowledge were more delay than tuberculosis patients who had good knowledge. This finding might be suggested that influence of patient awareness and understanding of tuberculosis symptoms, transmission, severity and treatment-seeking behaviors on the timing of diagnosis. Poor knowledge about tuberculosis may lead to underestimation of symptoms, delayed healthcare-seeking actions, and an overall lack of urgency in accessing medical care, ultimately contributing to diagnosis delays. The reason for this outcome might be patients who have poor knowledge of TB believe that TB is acquired from evil and they seek traditional healers or religious leaders to be freed from evil spirits. In the context of this study the majority of patients believed that their illness and symptoms will disappear by itself. This might be due to study participants with good knowledge about tuberculosis might have better inclination for early diagnose. This means having good knowledge of tuberculosis means of transmission, prevention diagnosis & treatment option are vital for early diagnosis of tuberculosis. This result supports the finding of consistent studies carried out in public health facility Gamo zone in Ethiopia the extent of knowledge regarding tuberculosis was significant factors for patient delays in diagnosis of tuberculosis (45). The study supports and inline this finding in Somali pastoralist region patients who had poor knowledge of tuberculosis symptoms, thought symptoms significant factors for delays in diagnosis tuberculosis(65). This finding is consistent with the study in north Shewa zone(27). Studies similar with this study carried out in Oromia Arsi zone respondents who have poor knowledge of TB delayed 2.7 more time than those patients who have the knowledge (29). Other study supports this finding tuberculosis patients in Central Myanmar showed that low level of knowledge significant factors to the delay in tuberculosis diagnosis (80).

## Conclusion

Diagnosis delay is a major challenge to eliminate tuberculosis by breaking the chain of transmission. This study can be attributed to factors including place of residence, knowledge about tuberculosis, initial actions upon symptom onset, and distance to health institutions. Recognizing these determinants helps in identifying and addressing barriers inhibiting early diagnosis, potentially improving healthcare access and promoting more timely diagnosis and treatment for tuberculosis patients.

## RECOMMANDATIONS

Based on the major findings of the study, recommendations for Metekele zone office and other tuberculosis program partners include addressing significant factors contributing to diagnosis delays through community-based interventions, targeted health education programs, strengthening referral systems, and improving access to healthcare services. Specific suggestions involve implementing community-based awareness campaigns, developing targeted health education programs for high-risk populations, improving referral systems, utilizing mobile health services for remote communities, training community health workers, and exploring telemedicine options. By implementing these evidence-based recommendations, delays in tuberculosis diagnosis can be reduced, ensuring timely access to proper healthcare services for affected individuals and ultimately improving overall tuberculosis control efforts in the region.

## Data Availability

Data used for this work is available at co-author

## Notes

### Competing Interest Statement

The authors have declared no competing interest.

